# A cross-sectional survey of burnout in a sample of resident physicians in Sudan

**DOI:** 10.1101/2021.11.25.21266880

**Authors:** Yasir Ahmed Mohamed Elhadi, Abdelmuniem Ahmed, Elhadi B. Salih, Osman S. Abdelhamid, Mohamed Hayder Hamid Ahmed, Noha Ahmed El Dabbah

## Abstract

**Background:** Resident physicians in Sudan suffer multiple physical and psychological stressors. Nevertheless, the prevalence of burnout among this critical population remains unknown. The purpose of this study was to estimate the prevalence of burnout and its related factors in a sample of Sudanese resident physicians.

**Methods:** A cross-sectional design was used to assess burnout among resident physicians at the teaching hospitals of Wad-Medani, Gezira state in Sudan. All resident physicians across nine hospitals were asked to join the study. The Arabic version of Maslach Burnout Inventory was distributed to respondents from July to October 2021.

**Results:** Out of 300 resident physicians 69.3% responded. The study population had a mean age of 29.99 ± 3.01 years, more than half were females (56.7%) and single (59.6%). In total, 86.1% met the criteria for burnout in at least one dimension, and 13.9 % in all three dimensions.70.7% suffer from a high level of burnout on the dimension of emotional exhaustion (EE), while 44.2% reported a high level on depersonalization (DP) and 73.1% experienced a sense of decreased professional accomplishment (PA). There were significant differences in the levels of burnout, EE and DP among different specialties. Burnout syndrome was associated with working hours per single duty; with participants working for more than 24 hours had experienced higher levels of burnout, EE and DP.

**Conclusions:** High prevalence of burnout syndrome was found in this sample of resident physicians in Sudan. Stakeholders should urgently implement effective remedies to protect the mental health of resident physicians.

## Background

Burnout syndrome (BOS) was first described by Freudenberg in 1974 [1]. In the 11^th^ revision of the International Classification of Diseases, BOS has been classified as an occupational phenomenon. It was defined as “A syndrome regarded stemming from continuous work stress that has not been properly managed”[2]. Burnout is characterized by three sub-dimensions; high emotional exhaustion, an increase in the mental distance from the profession; high depersonalization, a skepticism about the career; and a sense of decreased professional achievement [3].

Evidence suggests that healthcare workers suffer from high levels of BOS [4–6] with trainee physicians being at increased risk of BOS compared to other healthcare workers [7, 8]. Among trainee physicians, several occupational and individual factors have been recognized. However, it is uncertain which factors are the most significant in promoting the development of BOS [9].Work-related factors vary among different medical departments, which might suggest that there are certain fundamental disparities in working environments that can be linked to BOS among these populations [9]. BOS also appears to be influenced by payment models, with physicians who only receive incentive or performance-based pay have considerably higher burnout rates than salary paid physicians. [10, 11] BOS is influenced by the organizational context such as negative leadership behaviors and inadequate interprofessional collaboration, as well as possibilities for growth and social support for physicians [12]. BOS is also linked to poor work conditions, increased work demands, work-life imbalance, postgraduate training requirements interfering with personal life, and lack of senior support[9]. physicians’ burnout is not only financially costly, but also has a slew of other consequences, leaving Medicine is a clear example of such consequences [13]. In addition, depression, marital complications, medical errors, substance abuse, and suicidal behavior might occur [14]. Such consequences have detrimental impacts on healthcare organizations, physicians, and the quality of patient care provided [13].

The Sudan Medical Specialization Board (SMSB) is the professional training organization responsible for managing and delivering medical and health specialized programs in Sudan [15]. Sudan has been struggling to coordinate between human resources for health policies and the overall health planning. Because of the lack of coordination between the health authorities –represented by the Federal Ministry of Health- and medical training sector – represented in SMSB – there has been an imbalance between the training and production of health professionals in certain professions [16]. In addition, the mass migration of qualified health workers due to the economic crisis has left Sudan with a severe shortage in terms of qualified trainers. Moreover, the privatization of the health sector during the previous regime in Sudan has made the remaining qualified practitioners much less available for the education and training in teaching hospitals, which in turn affected the quality of medical training [17].

Despite being founded in 1995, the SMSB has just started paying less than 100 $ per month salary to resident physicians in 2021. At the same time, doctors accepted by training programs have to pay annual training fees to the SMSB, posing extreme financial burden on resident physicians and forces them to work extra-hours in private hospitals or clinics to cover their daily living expenses. This in turn affects their training quality and the time they have for academic achievement, which pose further risk for stress and burnout [18]. The training period ranges from four to seven years depending on the chosen specialty. For a considerable portion of the training period, a resident physician would have to work in other state hospitals in Sudan, away from their homes and families due to the SMSB random distribution of trainees. This scenario suggests extreme psychological and mental health impacts among resident physicians in Sudan that warrants urgent investigation to propose effective remedies. The purpose of this study was to estimate the prevalence rate of burnout and its associated factors in a sample of Sudanese resident physicians.

## Material and Methods

### Study design and setting

A cross-sectional design was used for this study. This survey was conducted at the teaching hospitals of Wad-Medani district, Gezira state, Southeast Sudan. There are nine teaching hospitals in Wad-Medani covering nine medical specialties, serving more than 3 million Sudanese population and affiliated with the University of Gezira-Faculty of Medicine, were selected for this study. The 300 resident physicians at dermatology, general surgery, pediatrics, obstetrics and gynecology, psychiatry, ear nose and throat, oncology, urology, and internal medicine departments, were approached and invited to participate in the present study.

### Data collection procedures

Data were collected from July to October 2021 through self-administered questionnaire that was comprised of two sections. The first section included the informed consent and other items related to sociodemographic and work-related attributes of participants including age, sex, marital status, specialty, and working hours/duty.

The second section contained the Arabic validated version of the Maslach Burnout Inventory Human Services Survey (MBI-HSS) [19], used after obtaining the required permissions from *Mind Garden*, Inc., an independent publisher of tools and instruments for psychological assessments of burnout, anxiety and leadership among others [20]. The MBI-HSS is the most accepted and widely used instrument for measuring burnout syndrome [21]. It consists of 22 items that investigate the three dimensions of BOS; nine questions for emotional exhaustion (EE); five questions for depersonalization (DP); and eight question for personal achievement (PA). Each item was ranked on a seven-point frequency rating scale, ranging from never (score 0) to every day (score 6). Higher scores on the EE and DP subscales were associated with higher levels of burnout, whereas high level of PA was associated with lower levels of burnout.

### Search strategy

In a similar fashion to other cross-sectional studies of burnout among healthcare workers [22], we performed online data search to compare burnout rates between resident physicians in Sudan and other countries. We searched PubMed database for similar cross-specialty studies of burnout among resident physicians published between 2000-2021. We excluded studies reporting burnout rate among single specialty resident physicians. The search results were evaluated by the authors and relevant studies were included and extracted on an excel sheet. The reported burnout rates in each included study were compared against the results of the current study.

### Data management and statistical analysis

Data were analyzed using IBM SPSS software package version 20.0. The Kolmogorov-Smirnov test was employed to ensure the normal distribution of variables. The three subscales for measuring burnout were categorized according to the scoring system of MBI-HSS [3] The Cronbach Alpha for the MBI-HSS in this study was 0.79 indicating the high reliability of the overall measurement. For normally distributed quantitative variables, the student t-test was used to compare two groups, while ANOVA was used to compare more than two groups. Significance of the results obtained was judged at the 5% level of alpha error.

### Ethical considerations

The current study was performed in accordance with research ethical standards and guidelines. The study was approved by the Health Sector Ethical Review Committee, University of Gezira (IRB approval No: 00036-21). Anonymity and confidentiality of data were guaranteed. A written consent to participate was included in the data collection tool and obtained from all participants. The right to withdraw from the study at any time was clearly stated prior to the start of the study.

## Results

The sociodemographic and work-related characteristics of respondents are shown in Table 1. A total of 208 resident physicians with mean age of 29.99 ± 3.01 years participated in the present study (69.3% response rate). More than half of respondents were females (56.7%), single (59.6%) and senior resident physicians (50.5%). Most of participants were medicine specialty residents (29.8%). More than one third reported working for more than 24 hours per single duty (32.7%) (Table 1).

**Table (1):** Sociodemographic and work-related characteristics of respondents.

| **Socio demographic characteristics** | **No** | **%** |
| --- | --- | --- |
| **Age (years)** |  |  |
| <30 | 104 | 50 |
| 30 – <35 | 84 | 40.4 |
| ≥35 | 20 | 9.6 |
| Mean ± Standard Deviation = $29.99 \pm 3.01$ | | |
| **Gender** |  |  |
| Male | 90 | 43.3 |
| Female | 118 | 56.7 |
| **Marital status** |  |  |
| Single | 124 | 59.6 |
| Married | 81 | 38.9 |
| Divorced/Widowed | 3 | 1.4 |
| **Specialty** |  |  |
| Medicine | 62 | 29.8 |
| Pediatrics | 38 | 18.3 |
| Urology | 13 | 6.3 |
| Obstetrics & Gynecology | 26 | 12.5 |
| Surgery | 38 | 18.3 |
| ENT | 7 | 3.4 |
| Oncology | 9 | 4.3 |
| Dermatology | 13 | 6.3 |
| Psychiatry | 2 | 1 |
| **Years of residency experience** |  |  |
| <3 | 103 | 49.5 |
| ≥3 | 105 | 50.5 |
| Mean ± Standard Deviation = $2.46 \pm 1.05$ | | |
| **Working hours per duty** |  |  |
| 8 to 11 hours | 37 | 17.8 |
| 12 to 17 hours | 50 | 24 |
| 18 to 24 hours | 53 | 25.5 |
| More than 24 hours | 68 | 32.7 |

The prevalence of burnout syndrome and its dimensions among resident physicians are shown in Table 2. 70.7% of resident physicians suffer from a high level of burnout on the dimension of emotional exhaustion, while 44.2% reported high levels on depersonalization and 73.1% experienced a sense of decreased professional accomplishment. Overall, 86.1% met the criteria for burnout in at least one dimension, and 13.9 % in all three dimensions.

**Table (2):** Frequencies of resident physicians by the levels of burnout in the three dimensions.

| Burnout and Symptoms | Low | Moderate | High |
| --- | --- | --- | --- |
| Emotional exhaustion (EE)<sup>a</sup> | 28 (13.5%) | 33 (15.9%) | 147 (70.7%) |
| Depersonalization (DP)<sup>b</sup> | 52 (25.0%) | 64 (30.8%) | 92 (44.2%) |
| Personal accomplishment (PA)<sup>c</sup> | 18 (8.7%) | 38 (18.3%) | 152 (73.1%) |
| Overall burnout<sup>*</sup> | 4 (1.9%) | 175 (84.1%) | 29 (13.9%) |

<sup>a</sup> Score of $\leq 16$ denoted a low level, score 17 to 26 denoted a moderate level, and score $\geq 27$ denoted a high level
<sup>b</sup> Score of $\leq 6$ denoted a low level, 7 to 12 denoted a moderate level, and score $\geq 13$ denoted a high level.
<sup>c</sup> Score of $\geq 39$ indicated low level, 32 to 38 denoted a moderate level; $\leq 31$ denoted a high level.
<sup>\*</sup> Overall burnout = burnout in all three dimensions (low scores on the EE and DP subscales, with a high score on the PA subscale, denoted a low degree of overall burnout; while high scores on the EE and DP subscales, with a low score on the PA subscale, denoted a high degree of burnout.

Parameters associated with burnout are shown in table 3. There were significant differences in the levels of emotional exhaustion (EE), depersonalization (DP) and the overall burnout among different specialties, with the Pediatric resident physicians showed higher overall burnout [mean ± SD, 3.19 ± 0.86 (P<0.001)], higher EE [mean ± SD, 4.36 ± 1.26(P<0.001)], and higher DP [mean ± SD, 3.09 ± 1.32) (P<0.001)]. Burnout, EE and DP were significantly associated with the working hours per single duty; with physicians working for more than 24 hours per a single duty, has experienced higher levels of burnout, EE and DP (P<0.001). However, there was no significant differences exist in the level of professional accomplishment related to respondents’ sociodemographic and work-related characteristics. Additionally, there were no significant differences noted in the levels of burnout and its three dimensions among different age groups, gender, years of experience, or marital status of resident physicians (Table 3).

**Table (3):** Parameters associated with burnout syndrome and its dimensions among resident physicians in Sudan.

| Socio-demographic characteristics | N | Emotional exhaustion<br>Mean ± SD. | Depersonalization<br>Mean ± SD. | Personal accomplishment<br>Mean ± SD. | Overall burnout<sup>a</sup><br>Mean ± SD. |
| --- | --- | --- | --- | --- | --- |
| **Age (years)** |  |  |  |  |  |
| <30 | **104** | 3.90 ± 1.36 | 2.95 ± 1.31 | 4.06 ± 1.05 | 2.97 ± 0.83 |
| 30 – <35 | **84** | 3.80 ± 1.48 | 2.88 ± 1.28 | 4.28 ± 1.05 | 2.84 ± 0.98 |
| ≥35 | **20** | 3.32 ± 1.51 | 2.47 ± 1.30 | 4.29 ± 1.0 | 2.54 ± 1.10 |
| **F (p)** |  | **1.401 (0.249)** | **1.150 (0.319)** | **1.184 (0.308)** | **1.965 (0.143)** |
| **Gender** |  |  |  |  |  |
| Male | **90** | 3.70 ± 1.35 | 3.03 ± 1.15 | 4.16 ± 1.05 | 2.87 ± 0.90 |
| Female | **118** | 3.88 ± 1.48 | 2.76 ± 1.39 | 4.17 ± 1.04 | 2.88 ± 0.94 |
| **t (p)** |  | **0.916 (0.361)** | **1.517 (0.131)** | **0.077 (0.939)** | **0.075(0.940)** |
| **Specialty** |  |  |  |  |  |
| Medicine | **62** | 3.93 ± 1.23 | 3.05 ± 1.22 | 4.20 ± 1.07 | 2.95 ± 0.87 |
| Pediatric | **38** | 4.36 ± 1.26 | 3.09 ± 1.32 | 4.07 ± 0.97 | 3.19 ± 0.86 |
| Urology | **13** | 3.32 ± 1.34 | 2.66 ± 0.81 | 3.99 ± 0.96 | 2.69 ± 0.71 |
| OBS & Gynae | **26** | 4.27 ± 1.26 | 3.45 ± 1.26 | 4.48 ± 0.91 | 3.08 ± 0.86 |
| Surgery | **38** | 3.84 ± 1.46 | 2.89 ± 1.25 | 4.23 ± 1.20 | 2.87 ± 0.98 |
| ENT | **7** | 3.54 ± 0.90 | 2.46 ± 1.35 | 4.27 ± 0.60 | 2.64 ± 0.78 |
| Oncology | **9** | 3.68 ± 1.07 | 2.40 ± 1.23 | 3.72 ± 0.65 | 2.88 ± 0.80 |
| Dermatology | **13** | 1.46 ± 0.82 | 1.29 ± 0.76 | 4.13 ± 1.0 | 1.57 ± 0.65 |
| Psychiatry | **2** | 2.44 ± 3.14 | 1.0 ± 1.41 | 2.94 ± 3.09 | 2.34 ± 0.48 |
| **F (p)** |  | **7.494\* (<0.001\*)** | **4.727\* (<0.001\*)** | **0.965 (0.464)** | **4.820\* (<0.001\*)** |
| **Years of residency experience** |  |  |  |  |  |
| <3 | **103** | 3.73 ± 1.51 | 2.83 ± 1.35 | 4.09 ± 1.08 | 2.86 ± 0.97 |
| ≥3 | **105** | 3.87 ± 1.34 | 2.92 ± 1.25 | 4.24 ± 1.01 | 2.89 ± 0.88 |
| **t (p)** |  | **0.722 (0.471)** | **0.461 (0.645)** | **1.016 (0.311)** | **0.186(0.853)** |
| **Marital status** |  |  |  |  |  |
| Single | **124** | 3.87 ± 1.35 | 3.0 ± 1.26 | 4.10 ± 1.02 | 2.95 ± 0.83 |
| Married | **81** | 3.75 ± 1.50 | 2.73 ± 1.34 | 4.25 ± 1.09 | 2.79 ± 1.03 |
| Divorced/Widowed | **3** | 2.63 ± 2.29 | 1.73 ± 0.42 | 4.79 ± 0.56 | 1.91 ± 1.22 |
| **F (p)** |  | **1.199 (0.304)** | **2.262 (0.107)** | **1.027 (0.360)** | **2.455 (0.088)** |
| **Working hours per duty** |  |  |  |  |  |
| 8 to11 hours | **37** | 2.72 ± 1.61 | 1.98 ± 1.47 | 3.80 ± 1.14 | 2.36 ± 1.05 |
| 12 to 17 hours | **50** | 3.69 ± 1.13 | 2.77 ± 1.03 | 4.37 ± 0.84 | 2.73 ± 0.74 |
| 18 to 24 hours | **53** | 4.06 ± 1.30 | 3.07 ± 1.16 | 4.13 ± 0.98 | 3.04 ± 0.80 |
| More than 24 hours | **68** | 4.27 ± 1.30 | 3.29 ± 1.25 | 4.26 ± 1.13 | 3.13 ± 0.94 |
| **F (p)** |  | **11.979\* (<0.001\*)** | **9.733\* (<0.001\*)** | **2.372 (0.072)** | **7.067\* (<0.001\*)** |

t: Student t-test
F: F for ANOVA test
N: Number
p: p value for comparing between the different categories
\*: Statistically significant at $p \leq 0.05$
<sup>a</sup> Overall burnout indicate high scores for burnout in all three dimensions

### Comparison of burnout rates among resident physicians in Sudan and other countries

The database search has generated 720 potentially relevant articles. After title, abstract and full texts screening, only 11 studies were included. The size of studies ranged from 68 in USA to 3350 in Syria. All included studies have reported higher prevalence of burnout syndrome compared to the current study. However, resident physicians in Sudan experienced higher levels of emotional exhaustion and depersonalization compared to resident physicians in the USA, KSA, Taiwan, Nigeria and Canada (Table 4).

**Table (4):** Comparing the present study results with other studies evaluating burnout among residents.

| Author, Year | Country | Sample size | Burnout rates | Comparison against current study results |
| --- | --- | --- | --- | --- |
| Zis, 2014 [23] | Greece | 263 | 14.4% of the residents were found to experience burnout | Slightly higher prevalence rate of burnout |
| Abdulrahman, 2018 [24] | UAE | 302 | 75.5% moderate-to-high EE, 84% had high DP, and 74% had a low sense of PA. | Higher EE, DP and PA. |
| Afzal, 2010 [25] | USA | 115 | 34 % reported high EE, 30% reported high DP and 24% reported low PA | Lower EE, DP and PA |
| Agha, 2015 [26] | KSA | 96 | Overall, 88.5% experienced burnout, with high EE in 68.8%, high DP in 63.6% and low PA in 38.5%. | Higher prevalence rate of burnout, EE and DP. With lower PA. |
| AlDubai, 2013[27] | Malaysia | 205 | High EE was reported by 36.6% of the respondents. DP and PA were not reported. | Lower EE, |
| Ben Zid, 2018 [28] | Tunisia | 149 | 17.14% had a high level of burnout | Higher prevalence of burnout. |
| Chaukos, 2016 [29] | USA | 68 | 28 % of resident physicians had a high level of burnout. | Higher prevalence of burnout. |
| Chen, 2013 [30] | Taiwan | 809 | 13.1% reported high EE. 9.3% reported high DP, and 0.7% reported high level of decreased PA | Lower EE, DP and PA. |
| Cheng, 2020 [31] | □USA | □ 105 | 71% prevalence of burnout. | Higher prevalence rate of burnout. |
| Cristina-Nituica 2021 [32] | USA | 682 | 51% prevalence of burnout of burnout. | Higher prevalence rate of burnout. |
| Ofei-Dodoo, 2019 [33] | USA | 218 | 43% prevalence for burnout, 32% reported high EE, 31% high DP, and 22% high level of decreased PA. | Higher prevalence rate of burnout. Lower EE, DP and PA. |
| Ogundipe, 2013 [34] | Nigeria | 204 | 45.6% had a high level of EE, 57.8% had high DP and 61.8% with high level of decreased PA. | Lower EE, DP and PA. |
| Alhaffar, 2019 [35] | Syria | 3350 | 19.3% prevalence of burnout syndrome. | Higher prevalence of burnout. |
| Ferguson, 2020 [36] | Canada | 718 | 69.4%. prevalence of burnout, with 61.6% high EE, 47.8% high DP and 29.0% high level of decreased PA. | Higher prevalence of burnout. Lower EE, DP and PA. |

## Discussion

This was a cross-sectional study designed to assess the prevalence of burnout syndrome and its related factors in a sample of resident physicians in Sudan. The results of the current study showed 13.9% prevalence rate of burnout among the participants (Table 2). Similar to a study conducted among resident physicians in Greece, in which 14.4% of residents met the criteria for burnout [23]. Additionally, a cross sectional survey in Tunisia concluded that 17.14% of resident physicians had a high level of burnout [28]. Another study in Syria [35] reported 19.3% prevalence rate of burnout syndrome among resident physicians.

In comparing the reported prevalence rate of burnout to existing literature, most cross-sectional studies of burnout syndrome have showed higher prevalence of burnout syndrome among resident physicians in other countries, compared to the results of the current study in Sudan (presented in Table 4). This could be explained by that most of these studies had used the short and abbreviated version of The Maslach Burnout Inventory, which was reported to overestimate the prevalence of burnout syndrome among resident physicians [37]. More importantly, the prevalence rate of burnout reported in the present study results is similar to other studies of burnout among other clinicians and health personnel in Sudan [38, 39], lending support to our findings.

In the analysis of parameters linked to burnout syndrome and its dimensions, there were significant differences noted across specialties with pediatric trainee suffer higher levels (Table 3). In support to this finding many other studies showed that pediatric resident with a higher risk, and suffer higher levels, of burnout syndrome [40–44]. Additionally, burnout rate was positively associated with working hours per duty of resident physicians. This finding was supported by results of a study conducted among resident physicians in Saudi Arabia which established the positive association between the resident duty working hours and burnout syndrome[45]. Moreover, there were no significant differences in burnout rates related to sociodemographic factors residents, age groups, gender, years of residency experience marital status (Table 3). A similar findings was reported in previously research of burnout among resident physicians in South Africa [46] and USA[47].

## Limitations

The current study doesn’t assess neither the level of stress, awareness of coping strategies nor job satisfaction among resident physicians in Sudan to avoid a long questionnaire. Hence, the selection of explanatory variables wasn’t theoretically grounded. However, the study was able to provide preliminary evidence regarding prevalence rate of burnout syndrome among resident physicians in Sudan. Since the study was conducted in Gezira state only, due to lack of sufficient resources for a nationwide survey, the results of study cannot be generalized to resident physicians in other states of Sudan.

## Conclusions

The study revealed high prevalence of burnout syndrome among the sample of resident physicians in Sudan, with the pediatric registrars being especially vulnerable. There was a significant difference in the level of burnout according to respondents’ working hours per duty. More studies are required to investigate the level of stress, awareness level of coping strategies, and job satisfaction among resident physicians in Sudan and to examine the association of these factors with burnout syndrome.

## Data Availability

All data produced in the present work are contained in the manuscript

## Statements and Declarations

### Funding

None.

## Acknowledgments

We would like to acknowledge the contributions of Dr. Adrian Rabe, PhD and his assistance in the current study.

## Conflict of interest statement

Authors declare no conflict of interest.

## Authors contributions

Conceptualization: [Yasir Ahmed Mohamed Elhadi, and Abdelmuniem Ahmed]; Data curation, Formal analysis and investigation:[All authors], Writing - original draft preparation: [Yasir Ahmed Mohamed Elhadi, Elhadi B. Salih, Osman S. Abdelhamid, and Mohamed Hayder Hamid Ahmed)]; Writing - review and editing: [All authors]; Supervision: [Noha Ahmed El Dabbah].

## Notes

### Competing Interest Statement

The authors have declared no competing interest.

### Funding Statement

This study did not receive any funding

### Author Declarations

This study was approved by the Health Sector Ethical Review Committee, University of Gezira (IRB approval No: 00036-21). Anonymity and confidentiality of data were guaranteed. A written consent to participate was included in the data collection tool and obtained from all participants. The right to withdraw from the study at any time was clearly stated prior to the start of the study.

